## Supplemental Information for "Mosquito and human characteristics influence natural Anopheline biting behavior and *Plasmodium falciparum* transmission"

**Affiliations**
[1]Duke Global Health Institute, Duke University, Durham, NC, USA
[2]Academic Model Providing Access to Healthcare, Moi Teaching and Referral Hospital, Eldoret, Kenya
[3]Duke Global Inc, Nairobi, Kenya
[4]Division of Infectious Diseases, Duke University School of Medicine, Durham, NC, USA
[5]Departments of Statistics & Sociology, University of Washington, Seattle, WA, USA
[6]Department of Epidemiology, Johns Hopkins Bloomberg School of Public Health, Baltimore, MD, USA
[7]School of Public Health, College of Health Sciences, Moi University, Eldoret, Kenya
[8]School of Medicine, College of Health Sciences, Moi University, Eldoret, Kenya

### Supplemental Methods

#### *P. falciparum* csp amplicon deep sequencing

All *P. falciparum*-positive DBS and reared mosquito abdomens were genotyped across a variable segment of the parasite circumsporozoite gene (*csp*) as previously described (1). Dual-indexed libraries were sequenced on an Illumina MiSeq platform, sequencing reads were demultiplexed and quality-filtered as previously described, and haplotype inference was performed using DADA2 as implemented in BRAVA (https://github.com/duke-malaria-collaboratory/BRAVA) (1). Initial haplotypes were quality-filtered as follows: haplotype length of 288 bp, minimum read depth of 125, minimum read proportion in a sample of 0.01, and minimum ratio *k/j* where *k* and *j* are haplotypes in the same sample such that *k* is one nucleotide different from *j* of 0.125. The sequence data are available from NCBI (BioProject PRJNA1064031).

#### Multilevel model analysis

For the multilevel risk factor analysis, we included as risk factors: gender, categorized age as of February 28, 2021 (<5, 5-15, >15), whether the person slept under a net (taken from the most recent monthly survey before the date of a matched bite; missing data was filled in using the monthly survey after the bite), and whether the person was infected with *P. falciparum* (nearest DBS result between -30 to +7 days of the bite, except if RDT+ -14 to 0 days from bite then negative after receiving treatment). We included as adjustors the number of STR-typed mosquitoes collected in the household that night, transmission season (high transmission season was defined as March to August), number of household members, whether any household member tested positive for *P. falciparum* by RDT in the previous month, and number of people sleeping in the same sleeping space as the person. Sensitivity analyses were performed on the *P. falciparum* infection risk factor by restricting to samples where the nearest DBS result was between -14 to +7 days or -7 to +7 days, or where a person was considered positive only when the nearest DBS before and after the bite were positive. Identical models were used with data subsetted to (1) only individuals infected with *P. falciparum* to identify the proportion of onward transmission to mosquitoes contributed by each group of individuals, and (2) only infectious mosquitoes to estimate relative entomological inoculation rates (EIRs).

**Data structure for discrete choice models**

Suppose we have a household consisting of 8 members with the following characteristics:

| Member | Age category (yrs) | Gender | Slept under net | *P. falciparum* infection status |
|---|---|---|---|---|
| A | > 15 | Male | Yes | Negative |
| B | > 15 | Female | Yes | Negative |
| C | 5-15 | Female | Yes | Positive |
| D | 5-15 | Male | No | Positive |
| E | > 15 | Female | No | Negative |
| F | < 5 | Male | Yes | Positive |
| G | < 5 | Female | Yes | Negative |
| H | < 5 | Female | Yes | Negative |

Now suppose all 8 household members were sleeping in the household on a given night in which 3 mosquito bloodmeals matched to the following people:

| Mosquito ID | Mosquito species | Multisource | Mosquito *Pf* status | Member(s) bitten |
|---|---|---|---|---|
| M1 | *An. gambiae* | No | Positive | D |
| M2 | *An. funestus* | No | Negative | D |
| M3 | *An. funsetus* | Yes | Positive | B & F |

Then, the data structure for the weighted discrete choice model multivariate on mosquito characteristics and univariate on human gender would be as follows:

| Mosquito ID | Mosquito species | Multisource | Mosquito *Pf* status | Human Gender | Choice | Weight |
|---|---|---|---|---|---|---|
| M1 | *An. gambiae* | No | Positive | Female | 0 | 5/8 |
| M1 | *An. gambiae* | No | Positive | Male | 1 | 3/8 |
| M2 | *An. funestus* | No | Negative | Female | 0 | 5/8 |
| M2 | *An. funestus* | No | Negative | Male | 1 | 3/8 |
| M3a | *An. funestus* | Yes | Positive | Female | 1 | 5/8 |
| M3a | *An. funestus* | Yes | Positive | Male | 0 | 3/8 |
| M3b | *An. funestus* | Yes | Positive | Female | 0 | 5/8 |
| M3b | *An. funsetus* | Yes | Positive | Male | 1 | 3/8 |

To estimate the relative risk ratio of choosing a male host over a female host for each mosquito co-variate, we run the following using the mlogit R package:

```
mlogit(choice ~ 1 | multisource + moz_head_pf_status + species, chid.var =
"index_mosquito", alt.var = 'gender', weights = weight, data = gender)
```

**Multisource meals and sleeping spaces**

To assess whether human characteristics associated with mosquitoes taking multisource meals were driven by shared sleeping spaces, we first generated pairs of cohort members living in the same households and assessed the relative risk that DNA from household member pairs who share sleeping spaces appeared in multisource bloodmeals compared to pairs who did not share sleeping spaces. Similarly, we assessed the unadjusted relative risk of individuals sharing a sleeping space with someone else by gender and age group. Unadjusted measures of association, wald confidence intervals, and chi squared tests were determined using the epi.2by2 function (epiR v2.0.66 (2)).

**Supplemental figures**

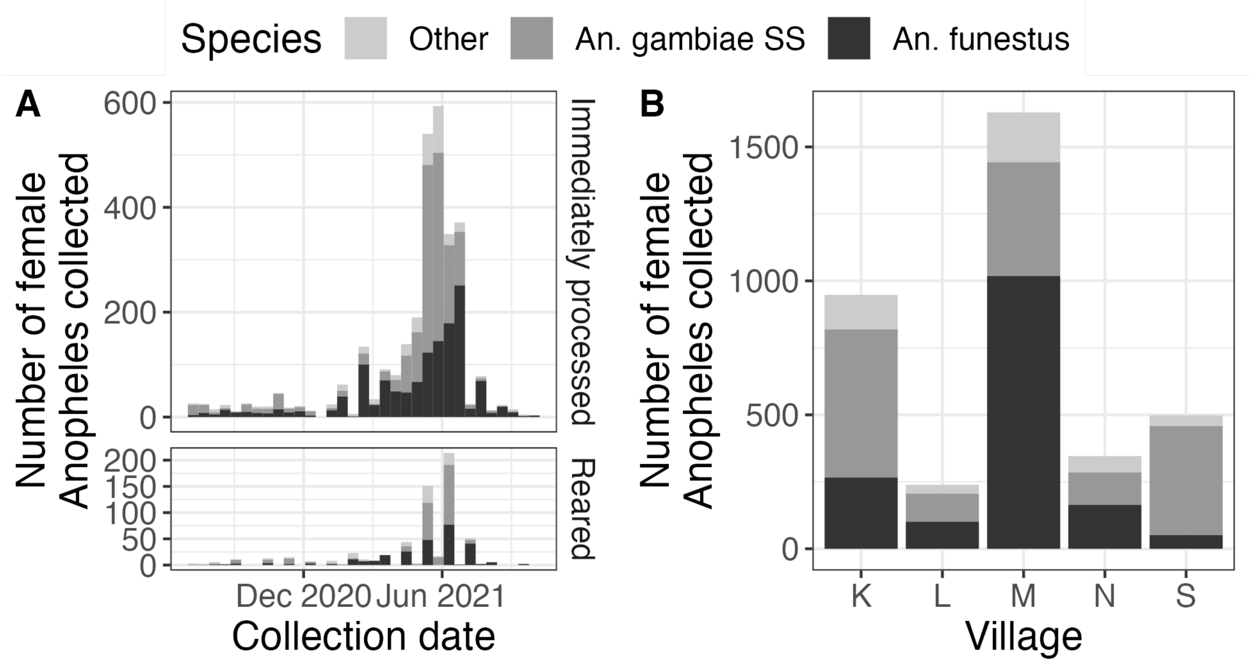


**Figure S1: Overview of female *Anopheles* collected.** (A) Number of female *Anopheles* collected over time, colored by species and faceted by immediately processed vs. reared. Bin width is 2-week intervals. (B) Number of female *Anopheles* collected by village, colored by species.

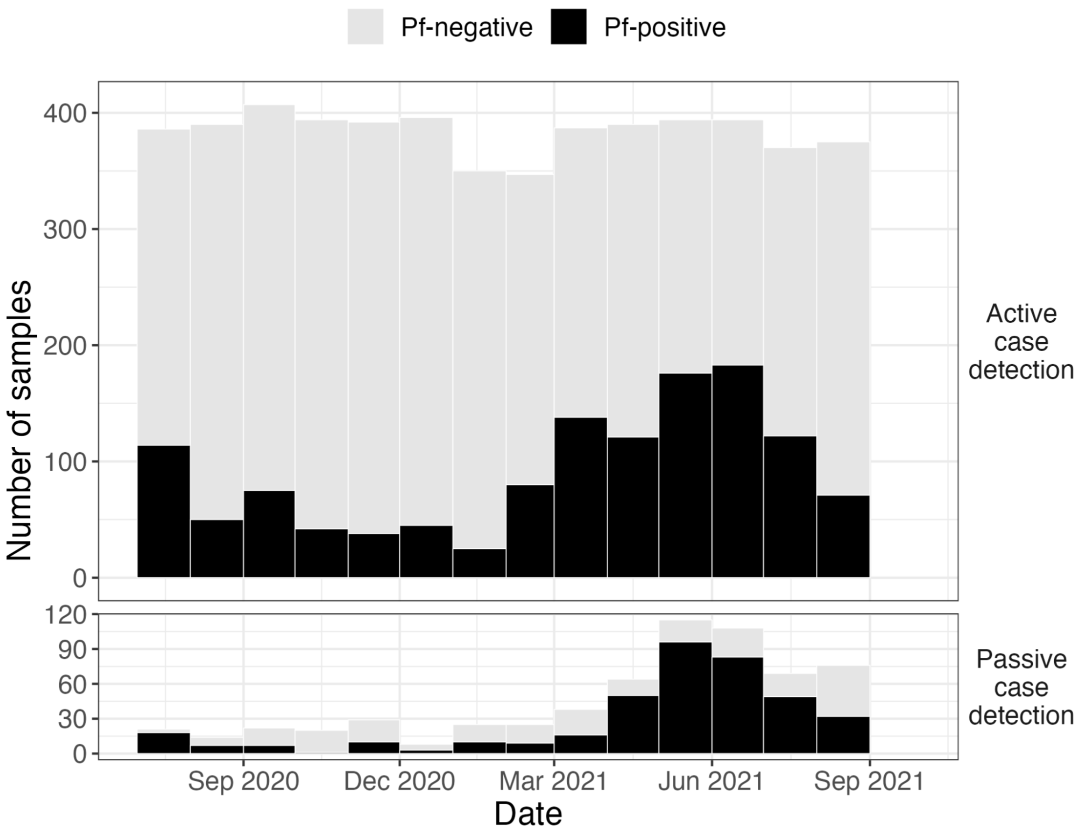


**Figure S2: Overview of human infections during the study period.**

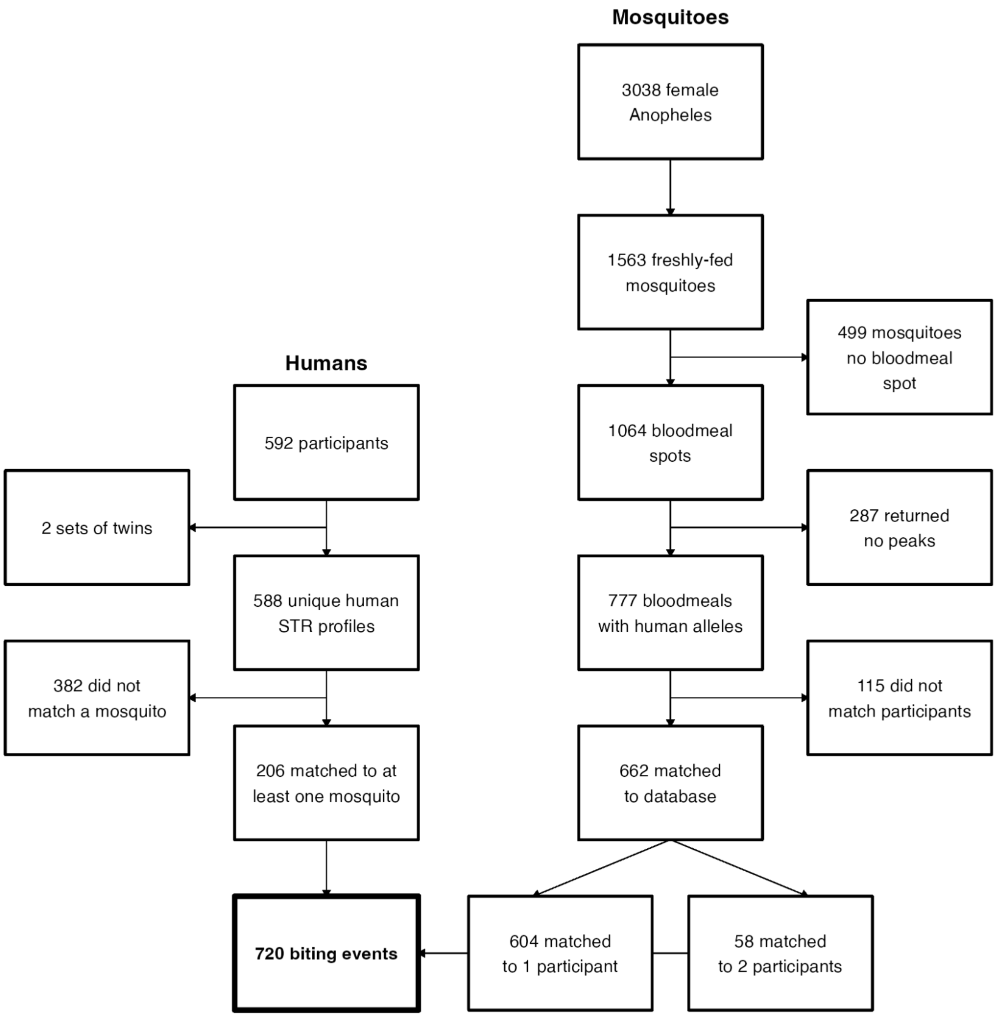


**Figure S3: Sample flowchart for immediately processed mosquitoes.**

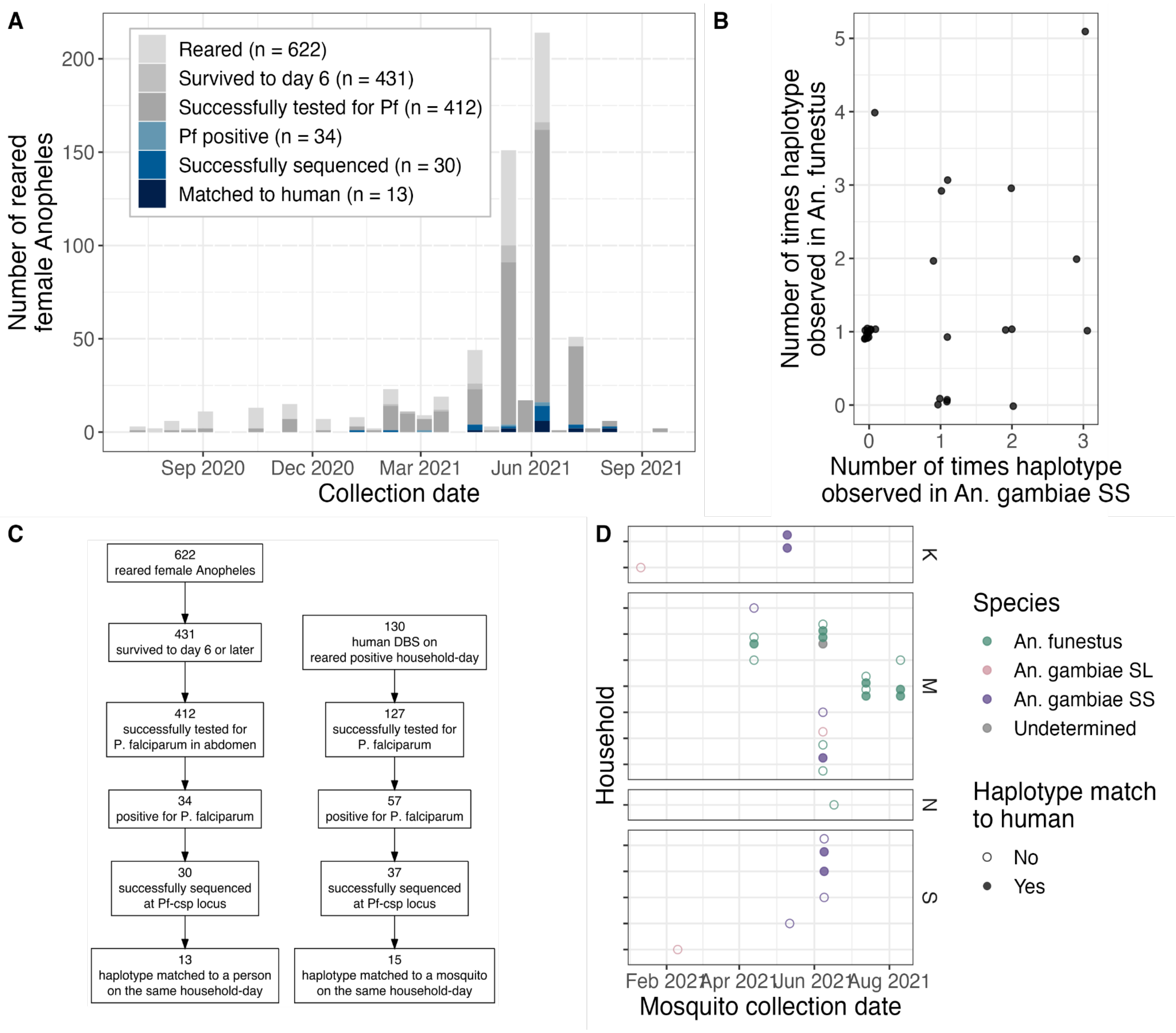


**Figure S4: Overview of reared mosquito spatiotemporal *P. falciparum* haplotype matching to humans.** (A) Number of reared processed mosquitoes (y) by collection date (x), colored by whether they were infected with *P. falciparum* and whether their haplotypes matched to an infected person from the same household-day. (B) Number of times a haplotype was observed in *An. gambiae SS* vs. *An. funestus*. (C) Sample flowchart for reared mosquitoes and humans from the household-dates where and when the mosquitoes were collected. (D) Mosquitoes that had a haplotype match to a human from the same household-day over time, colored by species.

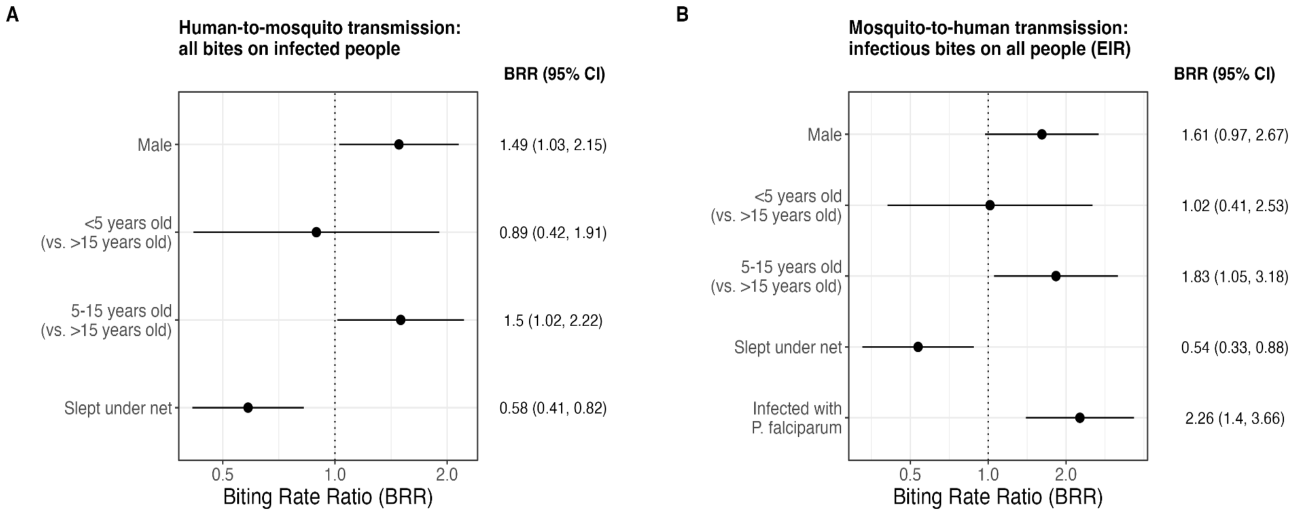


**Figure S5: Biting rate ratios from multilevel sub-models.** (A) Including only people infected with *P. falciparum.* (B) Including only infectious mosquitoes.

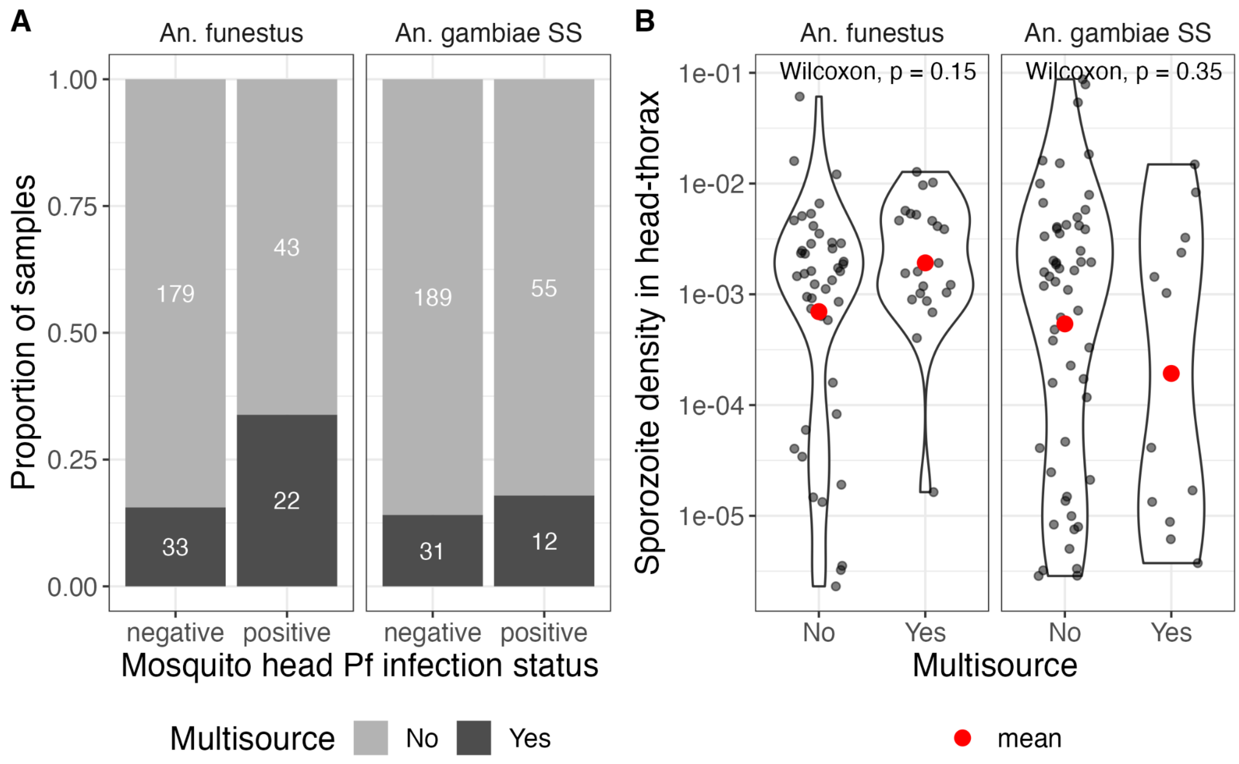


**Figure S6: Sporozoite density in head-thorax by multisource and species.**

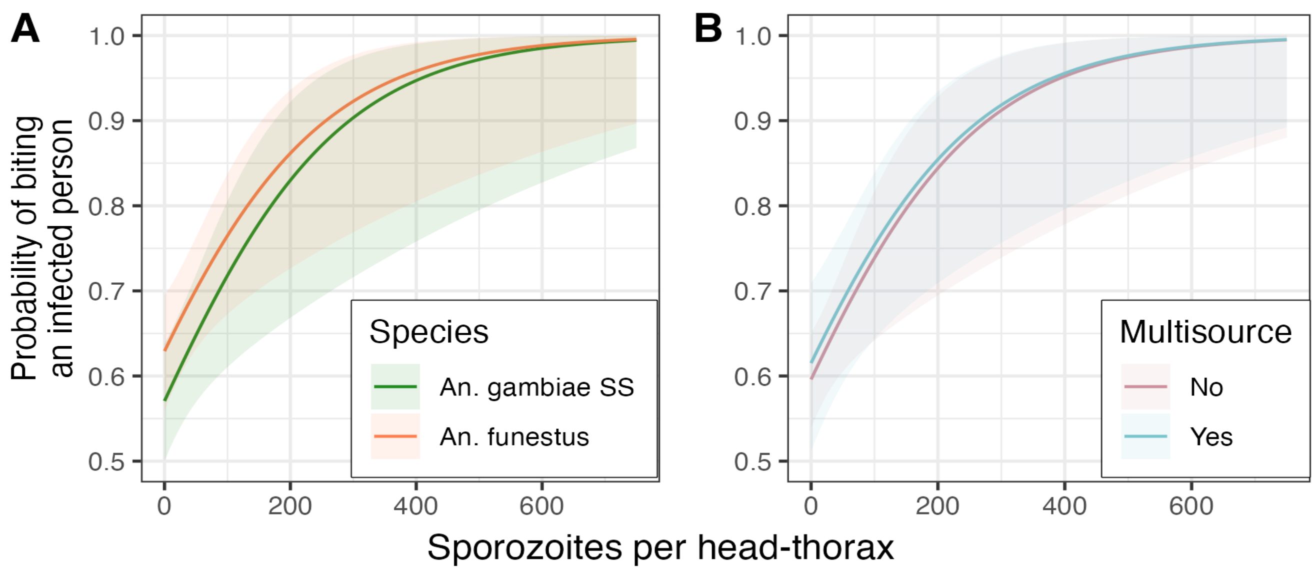


**Figure S7: Probability of biting an infected person as a function of sporozoite density.** Faceted by species (A) and multisource bloodmeal (B).

### Supplemental tables

#### Table S1: Risk factor analysis biting rate ratios

| Risk factor or adjustor | Biting Rate Ratio (95% CI) |
|---|---|
| Male | 1.68 (1.28-2.19) |
| <5 years old (vs. >15 years old) | 0.75 (0.44-1.29) |
| 5-15 years old (vs. >15 years old) | 1.49 (1.13-1.98) |
| Slept under net | 0.51 (0.40-0.65) |
| Infected with P. falciparum | 1.25 (1.01-1.55) |
| High transmission season | 1.31 (0.98-1.75) |
| Number of STR-typed mosquitoes in household | 1.13 (1.11-1.15) |
| Number of household members | 0.86 (0.81-0.90) |
| RDT+ household member in prior month | 0.86 (0.68-1.08) |
| Number of people in sleeping space | 0.90 (0.79-1.03) |

**Table S2: Sensitivity analyses decreasing the amount of time allowed between monthly visit and bite.**

| | Biting Rate Ratio (95% CI) | | | |
|---|---|---|---|---|
| **Model term** | **DBS window: -28 to +7 days** | **DBS window: -14 to +7 days** | **DBS window: -7 to +7 days** | **DBS+ before and after bite** |
| Male | 1.68 (1.28-2.20) | 1.58 (1.20-2.09) | 1.75 (1.24-2.46) | 1.76 (1.35-2.29) |
| <5 (vs. >15) years old | 0.75 (0.44-1.29) | 0.78 (0.45-1.36) | 0.28 (0.10-0.84) | 0.70 (0.41-1.19) |
| 5-15 (vs. >15) years old | 1.49 (1.12-1.98) | 1.43 (1.07-1.92) | 1.35 (0.95-1.92) | 1.37 (1.04-1.80) |
| Slept under net | 0.51 (0.40-0.65) | 0.46 (0.36-0.60) | 0.43 (0.31-0.60) | 0.51 (0.40-0.65) |
| Infected with P. falciparum | 1.25 (1.01-1.56) | 1.22 (0.97-1.53) | 1.22 (0.89-1.67) | 1.69 (1.32-2.17) |
| High transmission season | 1.30 (0.98-1.74) | 1.29 (0.93-1.78) | 1.33 (0.87-2.02) | 1.25 (0.94-1.66) |
| Number of STR-typed mosquitoes in household | 1.13 (1.11-1.15) | 1.13 (1.11-1.15) | 1.12 (1.09-1.14) | 1.13 (1.11-1.15) |
| Number of household members | 0.86 (0.81-0.90) | 0.85 (0.80-0.90) | 0.86 (0.80-0.93) | 0.86 (0.82-0.91) |
| RDT+ household member in prior month | 0.86 (0.68-1.09) | 0.95 (0.73-1.22) | 0.88 (0.63-1.22) | 0.79 (0.62-1.00) |
| Number of people in sleeping space | 0.90 (0.79-1.03) | 0.89 (0.78-1.03) | 0.93 (0.78-1.11) | 0.90 (0.79-1.04) |

#### Table S3: Choice models relative risk ratios

| Mosquito feature | | Relative Risk Ratio (95% CI) |
|---|---|---|
| | | Human *P. falciparum* infection: positive vs. negative (ref) |
| *An. funestus* vs. *An gambiae SS* (ref) | | 1.30 (0.87, 1.94) |
| Multisource vs. single-source bloodmeal (ref) | | 1.04 (0.64, 1.66) |
| Sporozoite positive vs. negative (ref) | | 2.76 (1.65, 4.61) |
| | | Slept under a net vs. did not sleep under a net (ref) |
| *An. funestus* vs. *An gambiae SS* (ref) | | 0.69 (0.41, 1.19) |
| Multisource vs. single-source bloodmeal (ref) | | 1.75 (0.97, 3.14) |
| Sporozoite positive vs. negative (ref) | | 0.45 (0.23, 0.86) |
| | | Human gender: male vs. female (ref) |
| *An. funestus* vs. *An gambiae SS* (ref) | | 1.19 (0.82, 1.72) |
| Multisource vs. single-source bloodmeal (ref) | | 0.50 (0.33, 0.77) |
| Sporozoite positive vs. negative (ref) | | 2.02 (1.28, 3.18) |
| | | Age vs > 15 years (ref) |
| | < 5 years | 5 - 15 years |
| *An. funestus* vs. *An gambiae SS* (ref) | 0.73 (0.32, 1.66) | 1.72 (1.17, 2.52) |
| Multisource vs. single-source bloodmeal (ref) | 4.10 (1.80, 9.35) | 1.45 (0.89, 2.35) |
| Sporozoite positive vs. negative (ref) | 2.69 (1.19, 6.09) | 2.21 (1.37, 3.59) |

#### Table S4: Choice model with continuous sporozoites (per 100 in the head-thorax)

| Mosquito feature | Relative Risk Ratio (95% CI) |
|---|---|
| | Human *P. falciparum* infection: positive vs. negative (ref) |
| *An. funestus* vs. *An gambiae SS* (ref) | 1.28 (0.86, 1.90) |
| Multisource vs. single-source bloodmeal (ref) | 1.08 (0.68, 1.73) |
| Sporozoite density in head-thorax (per 100) | 1.92 (1.23, 2.98) |
